# Histology-Derived Signatures Predict Recurrence Risk and Chemotherapy Benefit in Randomized Trials of Early Breast Cancer

**DOI:** 10.64898/2026.04.23.26351499

**Authors:** Frederick M. Howard, Anran Li, Sara Kochanny, Megan Sullivan, E Martin Flores, James Dolezal, Galina Khramtsova, Sasha Jain-Liu, Riley Medenwald, Poornima Saha, Cheng Fan, Linda McCart, Mark Watson, Lauren R. Teras, Clara Bodelon, Alpa Patel, W Fraser Symmans, Ann H. Partridge, Lisa Carey, Olufunmilayo I. Olopade, Daniel Stover, Charles Perou, Katharine Yao, Alexander T. Pearson, Dezheng Huo

**Affiliations:** Department of Medicine, University of Chicago, Chicago, IL, USA; Endeavor Health Cancer Institute, Evanston, IL, USA; Department of Pathology, Ingalls Memorial Hospital, Harvey, IL, USA; Geisinger Cancer Institute, Danville, PA, USA; Lineberger Comprehensive Cancer Center, University of North Carolina at Chapel Hill, Chapel Hill, NC, USA; The Ohio State University Comprehensive Cancer Center, Columbus, OH, USA; Department of Pathology and Immunology, Washington University School of Medicine, St. Louis, MO, USA; Department of Population Science, American Cancer Society, Atlanta, GA, USA; Department of Pathology, The University of Texas MD Anderson Cancer Center, Houston, TX, USA; Dana-Farber Cancer Institute, Boston, MA, USA; Department of Public Health Sciences, University of Chicago, Chicago, IL, USA

**Keywords:** breast cancer, digital pathology, predictive biomarkers

## Abstract

**Purpose:** To test whether histology-derived gene-expression signatures from routine hematoxylin and eosin slides are prognostic for recurrence and predictive of chemotherapy benefit in early breast cancer.

**Methods:** We conducted a multi-cohort study including CALGB 9344 (anthracycline ± paclitaxel), CALGB 9741 (standard vs dose-dense chemotherapy), a pooled Chicago real-world cohort, and the American Cancer Society (ACS) Cancer Prevention Studies-II and -3. Whole-slide images were processed with a previously described pipeline to generate 61 histology-derived signatures per patient. The primary endpoint was distant recurrence-free interval (DRFI), except in ACS, where breast cancer-specific survival was used. Secondary endpoints include distant recurrence-free survival (DRFS) and overall survival. The most prognostic signature in CALGB 9344, selected by Harrell’s C-index, was evaluated in additional cohorts. Signature-treatment interaction was assessed by likelihood-ratio tests. Multivariable Cox models incorporating age, tumor size, nodal status, estrogen/progesterone receptor status, and signature were fit in CALGB 9344 to improve risk stratification.

**Results:** A total of 7,170 patients were included across four cohorts. The top histology-derived signature in CALGB 9344 showed strong prognostic performance for 5-year DRFI (C-index 0.63) and performed well across validation cohorts (C-index 0.60, 0.70, and 0.62 in CALGB 9741, Chicago, and ACS, respectively). The strongest predictive signal for treatment benefit was observed for DRFS. High-risk cases identified by the signature demonstrated greater benefit from taxane in CALGB 9344 (adjusted hazard ratio [aHR] 0.76 for DRFS, 95% CI 0.66-0.88; interaction p=0.028), from dose-dense chemotherapy in CALGB 9741 (aHR 0.69, 95% CI 0.56-0.85; interaction p=0.039), and differential chemotherapy benefit in the Chicago cohort (aHR 0.84, 95% CI 0.59-1.21; interaction p=0.009). Combined clinical-histology models improved risk stratification and identified low-risk groups with a 2%-10% risk of distant recurrence or breast cancer death.

**Conclusion:** Histology-derived signatures from H&E images are broadly prognostic and, unlike clinical factors, may predict chemotherapy benefit.

**Highlights:**

- Histology-derived H&E signatures consistently predicted recurrence risk across randomized trials and real-world cohorts.
- A single cutoff of a low-risk histology signature predicted taxane benefit and dose-dense chemotherapy benefit.
- Combined clinical-histology models identified low-risk groups with 2%-10% risk of distant recurrence.

## Introduction

Seminal randomized trials from Cancer and Leukemia Group B (CALGB) demonstrated benefit of adding paclitaxel after doxorubicin-cyclophosphamide (AC) in node-positive disease^1^ (CALGB 9344/INT-0148) and of dose-dense chemotherapy (CALGB 9741)^2^, establishing the chemotherapy backbone for early breast cancer that is still the gold standard today. In parallel, multigene assays such as the 21-gene Recurrence Score (OncotypeDX) and the 70-gene signature (MammaPrint) demonstrated strong prognostic value, with subsequent trials demonstrating these assays predict chemotherapy benefit^3–6^. However, molecular assays remain variably accessible worldwide, can take weeks to perform, and may still misclassify risk and contribute to rising healthcare costs. Conversely, routine clinical workflows begin with hematoxylin and eosin (H&E) stained histology on all patients, and histology captures tumor architecture and immune contexture that reflect underlying molecular profiles.

Over the past decade, computational pathology has shown that deep learning can extract clinically relevant phenotypes directly from H&E. In breast cancer, image-based models have linked stromal and morphometric patterns to distant recurrence risk and survival, suggesting that histology encodes biologically meaningful information beyond conventional grading^7^. Deep learning models can achieve clinical-grade accuracy for hormone receptor status^8^ and HER2 amplification^9^, and can even accurately predict bulk gene expression^10,11^ or genomic risk signatures such as OncotypeDX^12,13^. Prognostic histology models have been developed on large patient cohorts by either directly predicting genomic risk scores^12^ or predicting patient prognosis^14,15^. However, prediction of genomic risk may not outperform standard molecular risk profiling. Moreover, although genomic risk assays are both prognostic and predictive, a more precise approach may uncover factors that directly predict chemotherapy benefit independent of prognosis. We previously developed Transformer-based hIstology-driven Gene Expression Regressor (TIGER), which accurately predicts 61 distinct histologic signatures based on gene pathways that concisely encapsulate diverse aspects of breast cancer biology^16^. In this earlier study we found several histology-derived signatures predicted response to specific therapies in the neoadjuvant setting. Thus, we assessed whether these signatures could predict prognosis and, more importantly, identify a group of patients who did not benefit from chemotherapy or treatment intensification in a large cohort of randomized adjuvant trials and real-world patients.

## Methods

### Study Design and Case Selection

We conducted a multi-cohort analysis to evaluate the prognostic and predictive utility of 61 histology-derived gene-expression signatures in early breast cancer. We included two randomized adjuvant chemotherapy trials: CALGB 9344 (AC ± paclitaxel)^1^ and CALGB 9741^2^ (standard-schedule vs dose-dense AC-T). Two real-world cohorts were used for additional validation: a pooled cohort from three Chicagoland hospitals (including University of Chicago, Endeavor Health Cancer Institute and Ingalls Memorial Hospital), and patients with hormone-receptor positive breast cancer from the American Cancer Society (ACS) Cancer Prevention Study-II^17^ and -3^18^. Inclusion criteria for the overall study were: invasive breast carcinoma; digitized H&E whole-slide images (WSI) from formalin-fixed paraffin-embedded slides; standard clinicopathologic covariates (age, tumor size, nodal status, ER and PR); documented chemotherapy regimen/arm where applicable; and availability of survival outcomes (**Figure 1**). The overall study was approved by the University of Chicago Institutional Review Board (IRB 22-0707). University of Chicago patients were prospectively consented under protocol IRB 16-352A from 2010 onward, whereas the Endeavor and Ingalls cohorts were retrospective and included diagnoses from 2010 to 2022. CPS-II and CPS-3 were approved by the Emory IRB (protocols IRB00045780 and IRB00059007). Included participants self-reported breast cancer on follow-up surveys, and were then asked for consent to obtain a medical record for verification and clinical details about the cancer and to request a tumor tissue sample from the hospital where it was stored.

**Figure 1.**
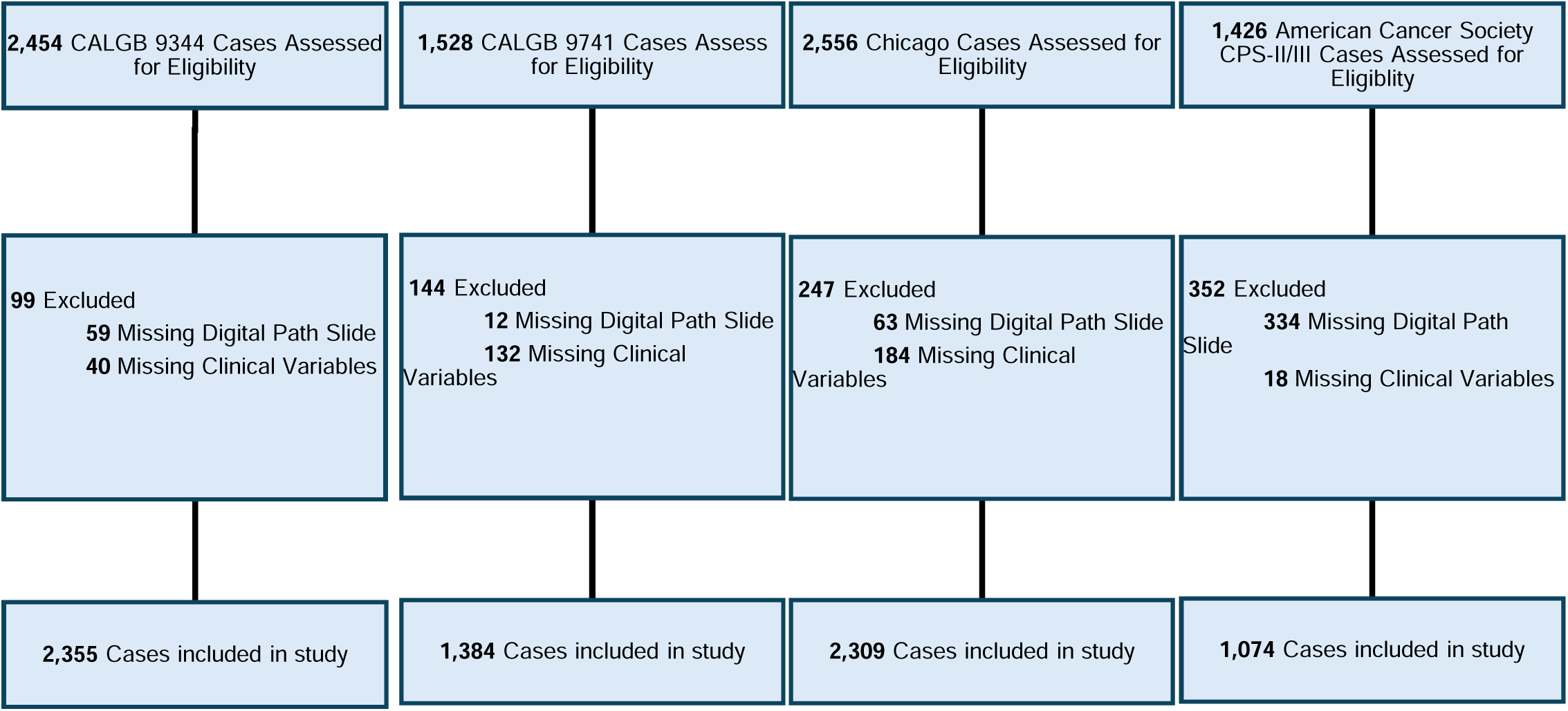
CONSORT Diagram for Included Patients Across Four Study Cohorts. **Abbreviations**: CALGB – Cancer and Leukemia Group B; CPS – Cancer Prevention Study.

### Image Extraction and Signature Generation

WSIs from the CALGB cohorts were scanned on a Leica Aperio AT2 at 20x, slides from the Chicago cohort were scanned on an Aperio AT2 at 40x, and slides from the ACS cohorts were scanned on a Hamamatsu scanner at 40x. Tiles of 224 pixels were extracted at an effective resolution of 10×. Regions with excess white or gray space were filtered out, and out-of-focus areas and pen markings were removed using Otsu thresholding and Gaussian blur-based filtering. The pretrained TIGER model was then applied (consisting of the UNI foundation model for feature extraction^19^ and transformer-based feature aggregation^20^ all implemented in the Slideflow pipeline^21^) to generate continuous scores for 61 breast cancer-relevant gene-expression signatures that correlate well (Pearson correlation > 0.5) with true signatures. For explainability, we generated model attention and prediction heatmaps and used the HistoXGAN pipeline to create representative synthetic images modified to increase or decrease model predictions^22^.

### Outcome Measures and Statistical Analysis

The primary endpoint was distant recurrence-free interval (DRFI) for all cohorts except ACS where DRFI was unavailable, and breast cancer-specific survival (BCSS) served as the primary endpoint. Secondary endpoints included distant recurrence-free survival (DRFS) and overall survival (OS). The most prognostic signature was identified in CALGB 9344 by Harrell’s concordance index (c-index) on the primary endpoint (DRFI) for further external testing in CALGB 9741, Chicago, and ACS cohorts and integration into multimodal models. Confidence intervals for c-index estimates were generated with 1,000 bootstrap resamples. Time-dependent AUC using 1-year windows was also used to assess prognostic accuracy for early vs late recurrence.

For interpretable multimodal recurrence prediction, we then built clinical-only and combined clinical-signature models in CALGB 9344, using continuous age, tumor size, and nodal status as well as binary estrogen receptor (ER) and progesterone receptor (PR) status. Model discrimination was summarized with c-index as well as hazard ratios (HRs) for medium-/ high-risk groups. Low risk was defined as the quantile achieving < 10% distant recurrence in CALGB 9344, and high risk defined as the quantile with > 50% distant recurrence. Separate clinical models were fit for DRFI, DRFS, and OS to better account for the association between age and non-breast cancer survival events. For ACS, the DRFS model was applied to the BCSS endpoint. A subpopulation treatment effect pattern plot (STEPP) was used to identify candidate cutoffs for taxane benefit in CALGB 9344. This threshold was then formally tested for chemotherapy intensification and benefit via likelihood ratio test, representing the interaction of signature category with taxane benefit in CALGB 9344, dose-dense chemotherapy benefit in CALGB 9741, and overall chemotherapy benefit in the Chicago cohort. Interaction models included the same clinical covariates described previously to allow uniform interpretation in the real-world cohorts where chemotherapy receipt was not randomized. We compared these results to other histology-derived signatures (Oncotype, MammaPrint), and stratification based on the combined clinicopathologic Cox model. All analyses were performed in Python 3.11 using the lifelines package for survival metrics. All statistical tests were two-sided and reported at the alpha = 0.05 significance level. False-discovery correction was applied when assessing the significance of c-index values across the 61 signatures for overall and early versus late recurrence.

## Results

### Prognostic Value of Signatures Across Cohorts

A total of 7,170 patients met inclusion criteria across four cohorts (**Table 1**): CALGB 9344 (*n* = 2,355), CALGB 9741 (*n* = 1,384), a pooled Chicago real-world cohort from three hospitals (University of Chicago, NorthShore, Ingalls; *n* = 2,309), and the American Cancer Society Cancer Prevention Study (ACS; *n* = 1,122). The CALGB trials enrolled younger patients (mean age 48.5 ± 9.8 in 9344; 50.6 ± 9.5 in 9741), with larger tumors (31.4 ± 18.9 mm; 28.8 ± 17.7 mm) and greater nodal involvement (5.0 ± 4.7; 4.3 ± 4.5 involved nodes). Hormone receptor status was mixed, with approximately 30% hormone receptor-negative cases in both CALGB trials. Conversely, the Chicago / ACS cohorts included older patients (mean age 58.3 ± 11.7 in Chicago; 61.2 ± 11.4 in ACS); smaller tumors (21.4 ± 18.3 mm; 17.4 ± 14.5 mm); lower nodal involvement (0.6 ± 2.1; 0.7 ± 2.0 involved nodes); and predominantly HR+ disease (10.8% and 0% hormone receptor-negative in Chicago / ACS respectively). Median follow-up was 144 months in CALGB 9344, 35 months in CALGB 9741, 68 months in Chicago, and 79 months in ACS.

**Table 1:** Baseline Clinicopathologic Characteristics of Cohorts Included in Analysis. **Abbreviations:** CALGB – Cancer and Leukemia Group B; ACS – American Cancer Society; HR – hormone receptor.

|  |  | **CALGB 9344** | **CALGB 9741** | **Chicago** | **ACS** |
| --- | --- | --- | --- | --- | --- |
| **n** |  | 2355 | 1384 | 2309 | 1122 |
| **Age, mean (SD)** |  | 48.5 (9.8) | 50.6 (9.5) | 58.3 (11.7) | 61.2 (11.4) |
| **Race / Ethnicity** | **Asian** | 34 (1.4) | 17 (1.2) | 138 (6.0) | 15 (1.3) |
|  | **Black** | 212 (9.0) | 148 (10.7) | 567 (24.6) | 14 (1.2) |
|  | **Hispanic** | 93 (3.9) | 50 (3.6) | 80 (3.5) | 34 (3.0) |
|  | **Other / Unknown** | 27 (1.1) | 18 (1.3) | 29 (1.3) | 11 (1.0) |
|  | **White** | 1989 (84.5) | 1151 (83.2) | 1495 (64.7) | 1048 (93.4) |
| **Menopausal Status** | **Postmenopausal** | 909 (38.6) | 716 (51.7) | 0 (0.0) | 831 (74.1) |
|  | **Premenopausal** | 1442 (61.2) | 659 (47.6) | 0 (0.0) | 253 (22.5) |
|  | **Unknown** | 4 (0.2) | 9 (0.7) | 2309 (100.0) | 38 (3.4) |
| **Tumor Size in mm, mean (SD)** |  | 31.4 (18.9) | 28.8 (17.7) | 21.4 (18.3) | 17.4 (14.5) |
| **Number of Involved nodes, mean (SD)** |  | 5.0 (4.7) | 4.3 (4.5) | 0.6 (2.1) | 0.7 (2.0) |
| **HR Status** | **Positive** | 1585 (67.3) | 946 (68.4) | 1950 (89.2) | 1122 (100.0) |
|  | **Negative** | 770 (32.7) | 438 (31.6) | 236 (10.8) | 0 (0.0) |
| **HER2 Status** | **Positive** | 0 (0.0) | 232 (16.8) | 160 (6.9) | 0 (0.0) |
|  | **Negative** | 0 (0.0) | 759 (54.8) | 1973 (85.4) | 850 (75.8) |
|  | **Unknown** | 2355 (100.0) | 393 (28.4) | 176 (7.6) | 272 (24.2) |
| **Grade, n (%)** | **1** | 0 (0.0) | 0 (0.0) | 417 (18.1) | 322 (28.7) |
|  | **2** | 0 (0.0) | 0 (0.0) | 1131 (49.0) | 564 (50.3) |
|  | **3** | 0 (0.0) | 0 (0.0) | 587 (25.4) | 191 (17.0) |
|  | **Unknown** | 2355 (100.0) | 1384 (100.0) | 174 (7.5) | 45 (4.0) |
| **Distant Recurrence, n (%)** | **No** | 1495 (63.5) | 1116 (80.6) | 2215 (95.9) | 0 (0.0) |
|  | **Yes** | 860 (36.5) | 268 (19.4) | 94 (4.1) | 0 (0.0) |
|  | **Unknown** | 0 (0.0) | 0 (0.0) | 0 (0.0) | 1122 (100.0) |
| **Survival, n (%)** | **Alive** | 1352 (57.4) | 1000 (72.3) | 2124 (92.0) | 976 (87.0) |
|  | **Dead** | 1003 (42.6) | 384 (27.7) | 185 (8.0) | 146 (13.0) |

In CALGB 9344, many of the 61 histology-derived gene-expression signatures demonstrated significant univariable associations with DRFI. The top-performing model in CALGB 9344 was based on the bronchioid gene signature. This signature was originally developed to identify a favorable-prognosis subtype of lung adenocarcinoma^23,24^, but has prognostic relevance in breast cancer. This signature was protective for recurrence, with the strongest numerical predictive value in CALGB 9344 for both overall (C-index 0.59 for inverse of signature) and early (< 5 year) recurrence (C-index 0.63). It also performed well in validation cohorts (C-index 0.60, 0.70, and 0.62 for early recurrence in CALGB 9741, Chicago, and early breast cancer death in ACS cohorts respectively). Examination of all 61 signature associations across trials revealed a time-dependent relationship with recurrence. The association of all 61 signatures with early (< 5 year) recurrence in CALGB 9344 and CALGB 9741 was highly consistent (**Figure 2A**), as was association with late (> 5 year) recurrence (**Figure 2B**). An S100 A9/A8 (calprotectin) histology signature^25^, which denotes a pro-inflammatory pathway that promotes immune evasion and chemoresistance, had the strongest predictive effect for late recurrence in CALGB 9344 (C-index of inverse signature 0.63). Time-dependent AUCs in CALGB 9344, computed in yearly intervals, confirmed dynamic risk relationships (**Figure 2C**). Signatures that appeared protective during early follow-up (years 1-5) including MammaPrint and the bronchioid signature reversed to associate with increased late risk, whereas several signatures associated with early high risk including OncotypeDX and the S100 A9/A8 signature were protective for late recurrence. Visual inspection of model attention and prediction heatmaps illustrated that predictions were largely derived from tumor-rich regions on slides (**Figure 3A/B**). Bronchioid signature predictions were associated with low grade histologic features, well-circumscribed tumors, and high differentiation / tubule formation (**Figure 3B**).

**Figure 2.**
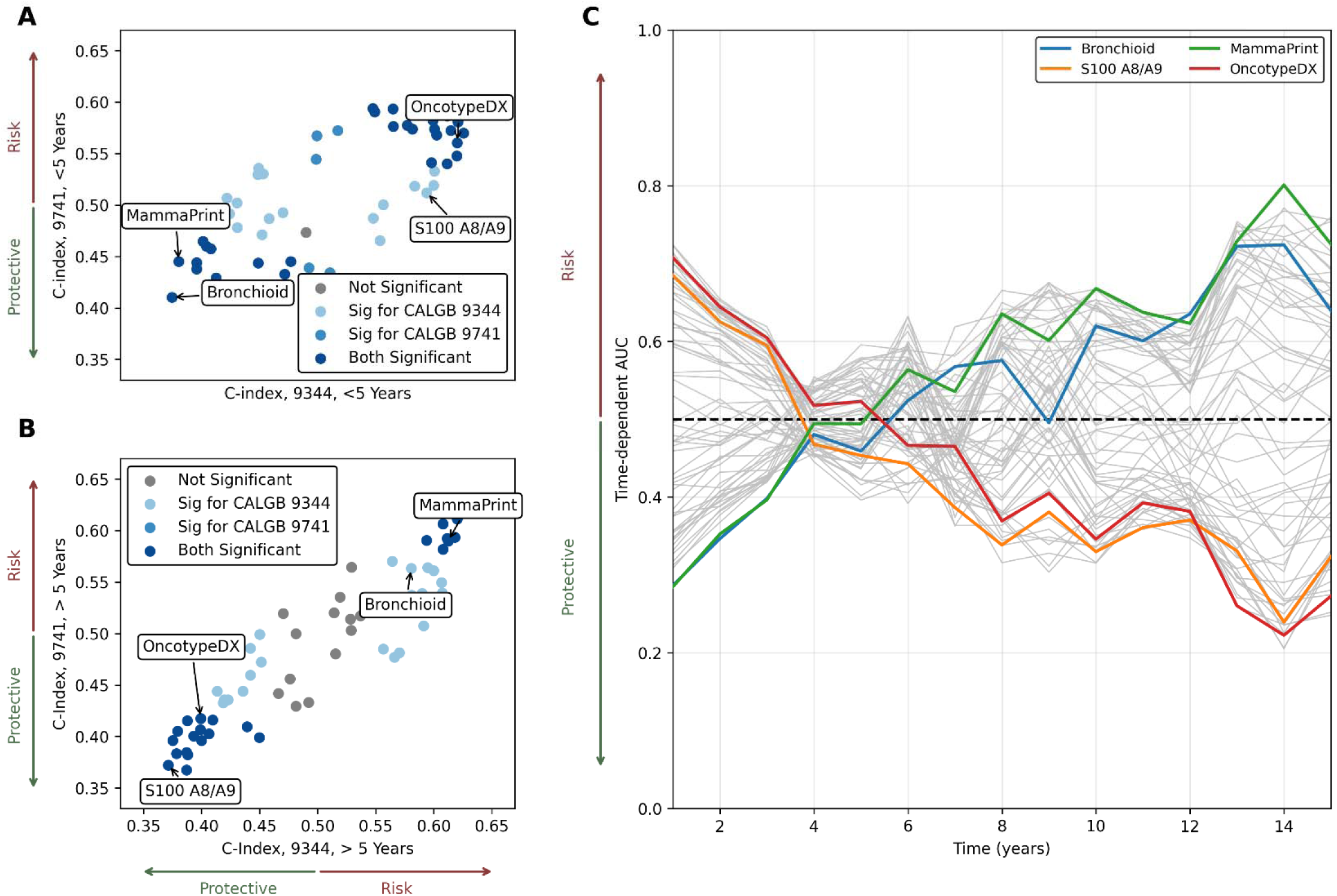
Histologic Features Have Inverse Associations with Early and Late Recurrence Risk. **A.** Correlation between concordance index (C-index) for prediction of early (< 5 year) recurrence in CALGB 9344 and CALGB 9741 demonstrates consistent risk / protective effects of histology-derived signatures for early recurrence. 1000x bootstrapping was used for significance testing as shown. **B.** Histology-derived signatures similarly have consistent predictive value for late recurrence in CALGB 9344 and 9741. **C.** Time-dependent areas under the ROC curve (AUCs) were estimated for 61 histology signatures in the CALGB 9344 trial using the window method, in which discrimination is calculated separately within successive yearly intervals rather than cumulatively. Each gray line represents one of the 61 signatures, while four signatures of interest are highlighted. Notably, most signatures that were protective during early follow-up (years 1-5) shifted to become predictive of increased risk in later years, and conversely, signatures that initially reflected higher risk showed protective associations at later times. **Abbreviations:** NS – not significant; Sig – significant; C-index – concordance index; CALGB – Cancer and Leukemia Group B; AUC – area under the receiver operating characteristic curve.

**Figure 3.**
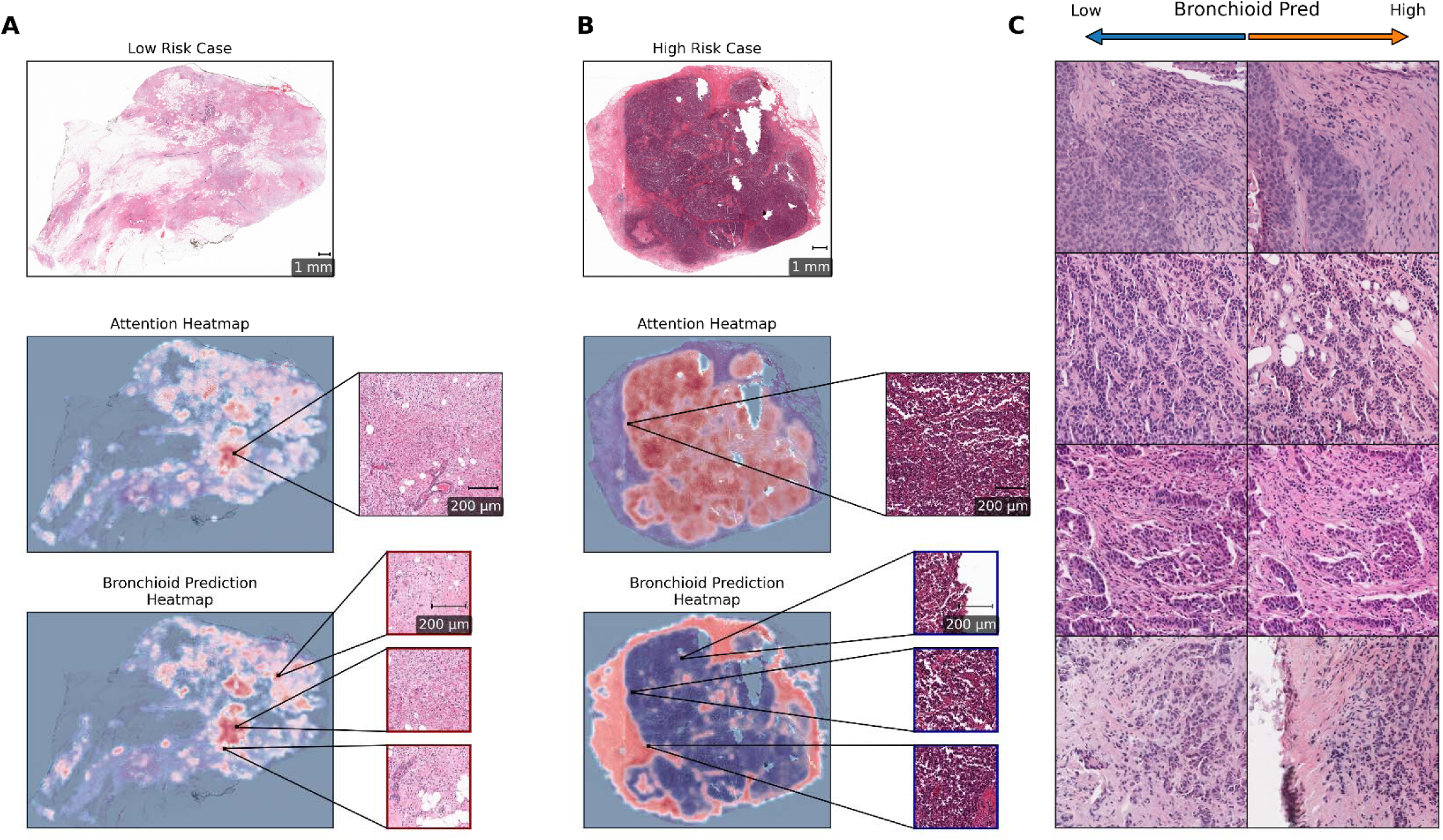
Composite Overview of Explainability of Bronchioid Signature Predictions. Shown are model predictions for a representative **A.** low-risk case and **B.** high-risk case. An attention heatmap illustrates slide areas that contribute to model predictions (with red indicating higher attention), which generally highlight tumor-rich regions of the slides. The prediction heatmap, with red indicating higher bronchioid signature prediction and therefore lower overall risk, illustrates the model signature prediction for each region of the slide. **C.** Digital histology images were generated based on representative cases, and then serially altered until model predictions were low / high using the HistoXGAN pipeline. High bronchioid signature (indicating low risk) was visually associated with lower histologic grade, less immune infiltrate, and higher differentiation / tubule formation.

### Reproducible Multimodal Prediction of Recurrence

Multivariable modeling of signatures and clinical variables confirmed violation of proportional hazards (**Supplemental Table 1**), and a pragmatic event restriction of 5 years was chosen for multivariable model fitting (to avoid combining predictors with opposing associations in early versus late recurrence). A multivariable clinical model comprising age, tumor size, nodal status, ER, and PR yielded a 5-year DRFI c-index of 0.67, which improved to 0.69 with the addition of the bronchioid signature (likelihood ratio test comparing models with and without the signature p < 0.001 for all endpoints). Similar improvements were seen with the addition of the signature for DRFI prediction in CALGB 9741 (c-index 0.66 to 0.68) and Chicago (c-index 0.74 to 0.78) and BCSS for ACS (c-index 0.60 to 0.67); as well as across other metrics in all cohorts (**Supplemental Table 2**). The bronchioid signature provided consistent independent predictive value with a protective effect in all multivariable models (**Supplemental Figure 1**). Performance was also consistent across clinicopathologic subgroups (**Supplemental Figure 2**). Using the combined model, thresholds were defined to identify low-risk disease (<10% risk of recurrence in CALGB 9344) and high-risk disease (>50% risk, **Supplemental Figure 3**). These thresholds identified low, medium, and high-risk groups with stepwise separation across cohorts (**Figure 4, Supplemental Tables 3 and 4**). The high risk group was associated with an increased rate of distant recurrence in the CALGB 9344 (HR 5.11, 95% CI 3.49-7.49), CALGB 9741 (HR 4.81, 95% CI 2.62-8.82), and Chicago cohorts (HR 18.31, 95% CI 8.88-37.73); with similar stepwise increases for distant recurrence-free survival (**Supplemental Figure 4**) and overall survival (**Supplemental Figure 5**). No ACS patients met the high-risk threshold, but the medium risk threshold also indicated an increased risk of breast cancer death (HR 2.96, 95% CI 1.70-5.17). The 8-year distant recurrence rate was 9.5% versus 50.3% in the low and high-risk groups for CALGB 9344, 10.3% versus 34.9% in CALGB 9741; and 1.9% versus 25.8% in the Chicago cohort. The 8-year rate of breast cancer death was 3.4% and 10.8% in the ACS low and medium-risk cohorts respectively.

**Figure 4.**
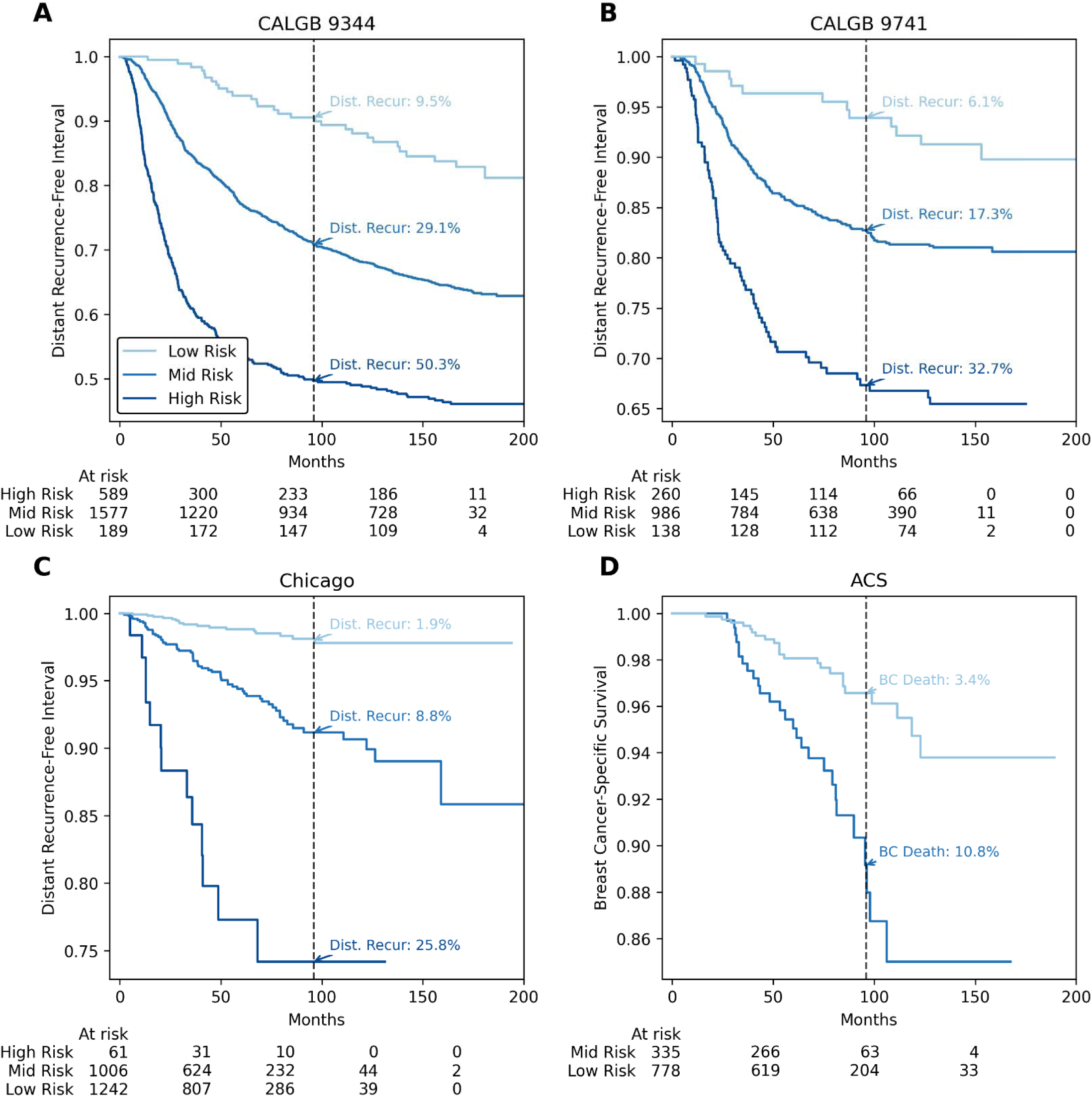
Multimodal Risk Stratification for Distant Recurrence / Survival Outcomes Across Clinical Trial and Real-World Cohorts. Curves show low, medium, and high-risk groups defined by thresholds fit in CALGB 9344 to identify patients with a risk of distant recurrence below 10% (low risk) or above 50% (high risk). These predictions are derived from Cox models that incorporate clinical covariates (ER status, PR status, tumor size, node status, age) and the histology-derived bronchioid signature. Shown are Kaplan-Meier estimates for distant-recurrence free interval in **A.** CALGB 9344; **B.** CALGB 9741; **C.** Chicago, and **D**. breast cancer-specific survival for ACS (as recurrence events were not captured in this cohort). The 8-year event rate is listed on each plot. **Abbreviations:** BC – Breast cancer; CALGB – Cancer and Leukemia Group B; ACS – American Cancer Society

### Predictive Value of Digital Histology Signatures for Treatment Benefit

We first assessed the predictive value of the bronchioid digital histology signature with respect to treatment benefit in the CALGB 9344 cohort. The STEPP analysis suggested that patients above the 70th percentile of the bronchioid signature derived minimal benefit from taxane chemotherapy, whereas greater benefit was observed below this threshold (**Figure 5A**). Conversely, at this cutoff, models fit on clinical features alone, or the clinicopathologic Cox model, did not have clear predictive value (**Supplemental Figure 6**). In an exploratory evaluation of all signatures, using a sigmoid transformation of signature value given the threshold-like association of the bronchioid signature with treatment benefit, we found that all predictive signatures were among the top 11 most prognostic signatures. Generally, histologic signatures that were most predictive of risk were able to distinguish patients who did not benefit from chemotherapy – with notable exceptions including the histologic OncotypeDX and MammaPrint models (**Supplemental Figure 7**).

**Figure 5.**
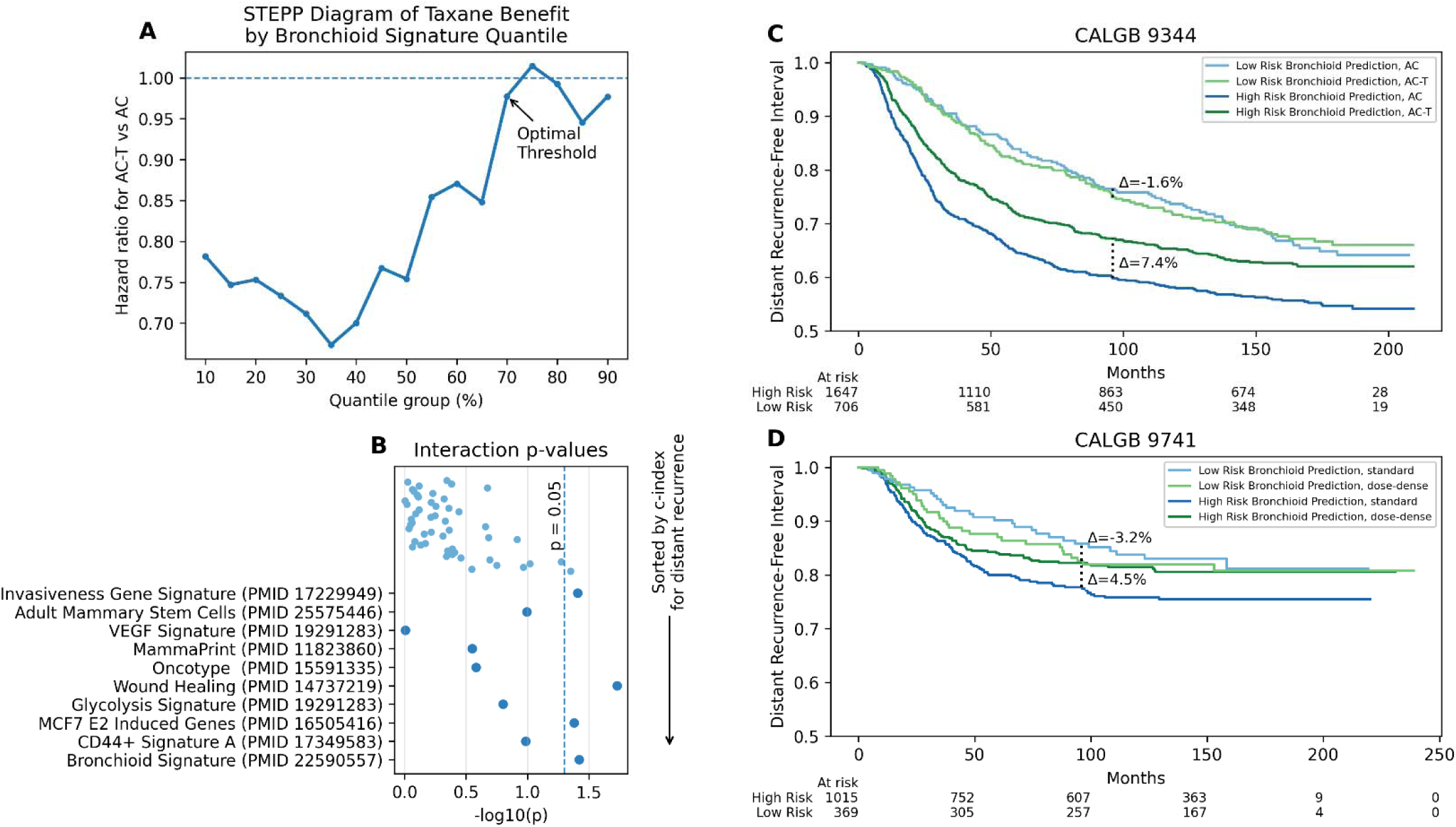
Histologic Signatures and Chemotherapy Benefit in Randomized Adjuvant Trials. **A**. Subpopulation treatment effect pattern plot illustrating the hazard ratio for taxane-containing chemotherapy (AC-T) versus AC across overlapping quantile-defined subgroups of the bronchioid histology signature score. A threshold of 70% appears to optimally select patients with the lowest benefit. **B**. Interaction analysis showing the significance of the treatment-by-signature interaction for candidate signatures in the CALGB 9344 cohort, ordered by concordance index for distant recurrence. For each signature, a Cox proportional hazards model for distant recurrence-free interval included treatment arm, a sigmoid-transformed signature term, and their interaction. **C.** Kaplan-Meier estimates of distant recurrence-free interval in CALGB 9344, stratified by predicted low- versus high-risk groups defined by the bronchioid signature (at the 70^th^ percentile) and randomized chemotherapy arm (AC vs AC-T). The 8-year landmark survival with annotated differences (Δ) in absolute survival between arms is listed. **D.** Kaplan-Meier estimates of distant recurrence-free interval in CALGB 9741 comparing standard-schedule versus dose-dense chemotherapy. Curves are stratified by predicted risk group (low vs high) using the same signature threshold. The 8-year landmark survival with annotated differences (Δ) in absolute survival between arms is listed. **Abbreviations:** STEPP – Subpopulation Treatment Effect Pattern Plot; CALGB – Cancer and Leukemia Group B; AC – doxorubicin and cyclophosphamide; T – paclitaxel.

As shown, the low-risk group of bronchioid prediction identifies patients with no benefit from adjuvant paclitaxel (aHR 1.02, 95% CI 0.81-1.27), whereas the higher risk group had significant benefit (aHR 0.78, 95% CI 0.67-0.91, interaction p = 0.136). This represented a 7.2% reduction of distant recurrence with taxane at 8 years in the high-risk group (**Figure 5C**). Similarly, significant benefit was seen in the high-risk group for DRFS (aHR 0.76, 95% CI 0.66-0.89; interaction p = 0.028) and OS (aHR 0.77, 95% CI 0.66-0.89, interaction p = 0.043) – but not in the low-risk groups for these outcomes. Furthermore, significant benefit in CALGB 9741 was seen for dose-dense vs standard-schedule treatment in the high-risk group for DRFI (aHR 0.74, 95% CI 0.58-0.95, interaction p = 0.123), representing a 4.2% reduction in distant recurrence with dose-dense chemotherapy at 8 years in the high-risk group (**Figure 5D**). Similarly, dose-dense treatment was associated with improved DRFS (aHR 0.69, 95% CI 0.56-0.85, interaction p = 0.039) and OS (aHR 0.71, 95% CI 0.57-0.89, interaction p = 0.079) only in the high-risk group. In the Chicago cohort, there was a trend towards improved DRFS with chemotherapy in the high-risk group (aHR 0.84, 95% CI 0.59-1.21) – with significant interaction based on signature group (p = 0.009). Consistent lack of treatment benefit was seen in low-risk subgroups of patients with hormone receptor-positive and -negative disease (**Supplemental Figure 8**), as well as other clinicopathologic subgroups (**Supplemental Figure 9**).

## Discussion

Here, we describe one of the first digital histology biomarkers to demonstrate both prognostic and treatment-predictive value in early breast cancer, with validation across over 7,000 breast cancer cases from clinical trials and real-world cohorts. Across multiple studies, the combined clinical and pathologic model developed here identified a low-risk group with a < 5% risk of distant recurrence / breast cancer death in the modern real-world cohorts, and a high-risk group with markedly worse outcomes. Additionally, we demonstrate that a pathologic risk signature alone, and not clinical or multimodal risk prediction, is predictive of treatment intensification, with significant interaction with respect to distant recurrence-free survival for taxane benefit, dose-dense chemotherapy benefit, and general chemotherapy benefit across three cohorts. Early work demonstrated that machine learning on digital pathology and clinical variables can predict genomic risk scores. Models such as hist2RNA linked slide morphology to expression of assay genes and PAM50-like signals, showing prognostic value in external tissue microarrays and multivariable survival analyses^26^. Subsequent studies scaled cohort size and began integrating machine learning approaches on both clinicopathologic factors^27,28^ and slide images^13^. Current approaches for risk prediction from digital images generally take one of two forms: (i) prediction of a genomic risk score from histology for clinical application and (ii) direct prediction of recurrence risk from histology.

Recent state of the art studies for predicting Oncotype score from histology include a multimodal model incorporating digital pathology and text pathology reports trained with data from Memorial Sloan Kettering, with Pearson correlations up to 0.60 on internal and external validation cohorts^29^. Another model, trained on data from TAILORx, achieved correlation coefficients on external cohorts of up to 0.72 using both clinical characteristics and H&E images^30^. Our OncotypeDX signature achieved comparable performance when compared with gene expression-derived Oncotype scores, demonstrating correlation coefficients of up to 0.70^16^. Importantly, although this Oncotype histologic signature was prognostic, it did not predict chemotherapy benefit in CALGB 9344. Previous studies have suggested that certain components of Oncotype (such as proliferation scores) may better predict chemotherapy benefit – for example, cases with high RS and PR negativity have less benefit from anthracyclines than PR positive cases, presumably due to a higher proliferation or other subscore driving the high RS^31^. Thus, training models to recapitulate Oncotype score from histology may not capture the precise histologic features that predict chemotherapy benefit, even if such models are prognostic. Alternatively, Oncotype may not accurately predict benefit from taxane chemotherapy, as suggested by a correlative analysis of NSABP B28^32^, and thus it may be expected that a digital histology Oncotype score fails to predict taxane benefit.

On the other hand, digital histology prognostic models are commercially available or in development that were originally trained to directly predict breast cancer outcomes^14,33^. However, as we show here, there are multiple key considerations for tuning such models to identify patients who derive little or no benefit from chemotherapy. First, we demonstrate that multimodal models, incorporating histology and clinical risk, may fail to identify patients who do not benefit from chemotherapy. Although they can identify a group with an overall low risk of distant relapse, clinical factors do not accurately distinguish the biologic features in breast cancer that predict benefit from chemotherapy. Similar findings were seen in an analysis of a multimodal model in NSABP B20 - although low-risk patients had lower recurrence risk, there was no interaction between either continuous or dichotomized multimodal AI risk scores and treatment benefit^34^. Additionally, training models on datasets with extended follow-up may conflate histologic features associated with early and late recurrence. Thus, in developing multimodal AI models, digital pathology estimates alone (as a surrogate for tumor biology) should be explicitly tested to answer whether chemotherapy offers any benefit, whereas aggregate multimodal risk estimates may help determine the magnitude of that benefit.

Our study has several important limitations. We selected the most predictive signature in CALGB 9344, which was a study that included multiple subtypes of breast cancer, patients received older endocrine therapy regimens, and HER2-positive patients did not receive HER2-directed therapy. Nonetheless, the consistent predictive and prognostic performance in modern, predominantly hormone receptor-positive cohorts suggest generalizability of this approach. These findings also support further evaluation of the predictive signature in HER2-positive and triple-negative disease, where biomarkers of treatment benefit remain limited. Interaction testing for DRFI did not reach statistical significance using the 70th-percentile cutoff. However, in cohorts such as CALGB 9741, the type of late recurrence (local vs distant) was not captured, which may have reduced power for DRFI-based analyses. In this context, the consistent significant interaction observed for DRFS across treatment settings supports the strength of the predictive signal. Conclusions regarding the predictive accuracy of true gene expression signatures cannot be drawn based on findings from our histology surrogates. Although these surrogates are moderate to strongly correlated with true signatures, there are features derived from gene expression that are poorly captured on histology. However, we have previously shown that these signatures are highly reproducible when captured from multiple slides from the same patient^16^, and thus can serve as a consistent risk predictor when applied to clinical samples.

## Conclusion

A comprehensive multimodal risk model based on histologic signatures can accurately predict recurrence risk, whereas a histology signature alone predicts patients who do not benefit from escalation of chemotherapy in multiple cohorts. Prediction of chemotherapy benefit should be grounded in tumor biology rather than clinical stage, and models trained to predict recurrence risk or emulate existing gene-expression assays should be carefully evaluated before clinical implementation.

## Acknowledgements

Cancer and Leukemia Group B (CALGB) is now part of the Alliance for Clinical Trials in Oncology, a National Clinical Trials Network cooperative group. Data and slides from studies CALGB 9344 and CALGB 9741 were obtained from the Alliance for Clinical Trials in Oncology, under Alliance data sharing agreement A152324. The authors express sincere appreciation to all Cancer Prevention Study-II and Cancer Prevention Study-3 participants, and to each member of the study and biospecimen management group. The authors would like to acknowledge the contribution to this study from central cancer registries supported through the Centers for Disease Control and Prevention’s National Program of Cancer Registries and cancer registries supported by the National Cancer Institute’s Surveillance Epidemiology and End Results Program. Through acceptance of this federal funding, NIH has been given a right to make the Author Accepted Manuscript publicly available in PubMed Central upon the Official Date of Publication, as defined by NIH. The content is solely the responsibility of the authors and does not necessarily represent the official views of the National Institutes of Health or the American Cancer Society or American Cancer Society -- Cancer Action Network.

This manuscript is the result of funding in whole or in part by the National Cancer Institute of the National Institutes of Health (NIH) under Award Numbers U10CA180821, and U24CA196171 (to the Alliance for Clinical Trials in Oncology), UG1CA233327, UG1CA189847, UG1CA233329, UG1CA233373, UG1CA233180, UG1CA233290, UG1CA233331, and UG1CA233339. Also supported in part by Bristol Myers Squibb (CALGB 9344).

This work was also supported by the following research grants:

National Cancer Institutes grant K08CA283261 (FMH)

Cancer Research Foundation grant (FMH)

Lynn Sage Breast Cancer Foundation (FMH)

Alliance Foundation Trials Special Projects Award (FMH)

Breast Cancer Research Foundation Next Generation Award (FMH)

National Cancer Institutes grant P20-CA233307 (OIO, DH)

Department of Defense grant BC211095P1 (FMH, ATP, DH)

National Institute of Dental and Craniofacial Research grant R56DE030958, (ATP)

National Cancer Institutes grant R01CA276652 (ATP)

European Commission Horizon grant 2021-SC1-BHC, (ATP)

Adenoid Cystic Carcinoma Research Foundation grant (ATP)

Cancer Research Foundation grant (ATP)

American Cancer Society grant (ATP)

National Cancer Institutes grant P50CA058223 (CMP)

Breast Cancer Research Foundation BCRF-23-127, (CMP)

Stand Up To Cancer grant (ATP)

## Data Availability

De-identified patient data from the CALGB 9344 and 9741 trials may be requested from Alliance for Clinical Trials in Oncology via if data are not publicly available. A formal review process includes verifying the availability of data, conducting a review of any existing agreements that may have implications for the project, and ensuring that any transfer is in compliance with the IRB. The investigator will be required to sign a data release form prior to transfer. Data from the Cancer Prevention Studies are available from the American Cancer Society by following the ACS Data Access Procedures (https://www.cancer.org/research/population-science/research-collaboration.html) for researchers who meet the criteria for access to confidential data. Please to inquire about access. Data from the Chicago cohort can be obtained via reasonable request to study authors. All code and trained models used for this analysis are available at https://github.com/fmhoward/TIGER.

---

**Supplemental Figure 1.**
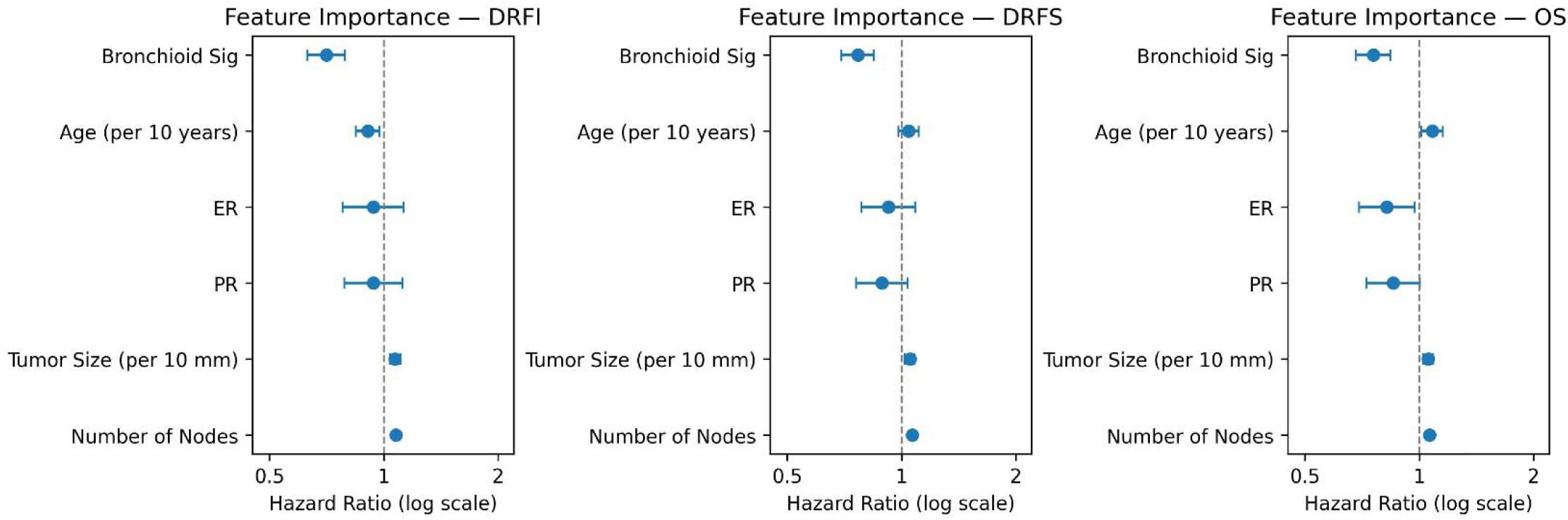
Feature Importance of Parameters of Multivariable Cox models Incorporating Clinical and Pathologic Factors from CALGB 9344. Hazard ratios with 95% confidence intervals are shown for distant recurrence-free interval, distant recurrence-free survival, and overall survival, using models trained in CALGB 9344 with follow-up capped at 5 years. **Abbreviations:** CALGB – Cancer and Leukemia Group B; DRFI – distant recurrence-free interval; DRFS – distant recurrence-free survival; OS – overall survival; ER – estrogen receptor; PR – progesterone receptor.

**Supplemental Figure 2.**
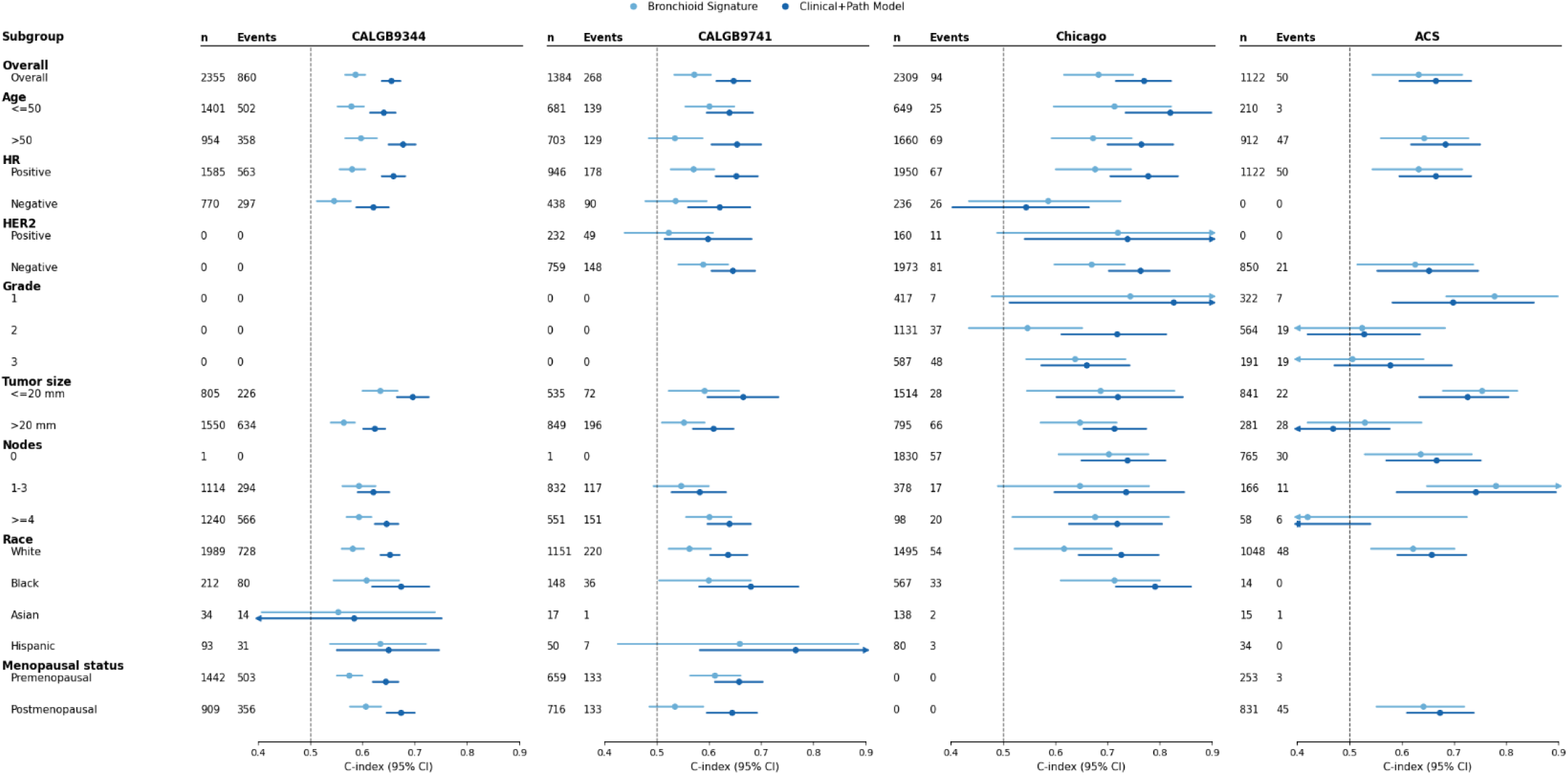
Prognostic Value of the Bronchioid Signature and Clinicopathologic Model Across Subgroups. Forest plots list C-index with 95% confidence intervals for the bronchioid signature and a clinicopathologic Cox model incorporating age, tumor size, nodal status, and estrogen/progesterone receptor expression. Prognostic performance is shown across clinicopathologic subgroups for distant recurrence-free interval in CALGB 9344, CALGB 9741, and Chicago cohorts, and breast cancer-specific survival in the ACS cohort. **Abbreviations:** CALGB – Cancer and Leukemia Group B; HR – hormone receptor; ACS – American Cancer Society.

**Supplemental Figure 3.**
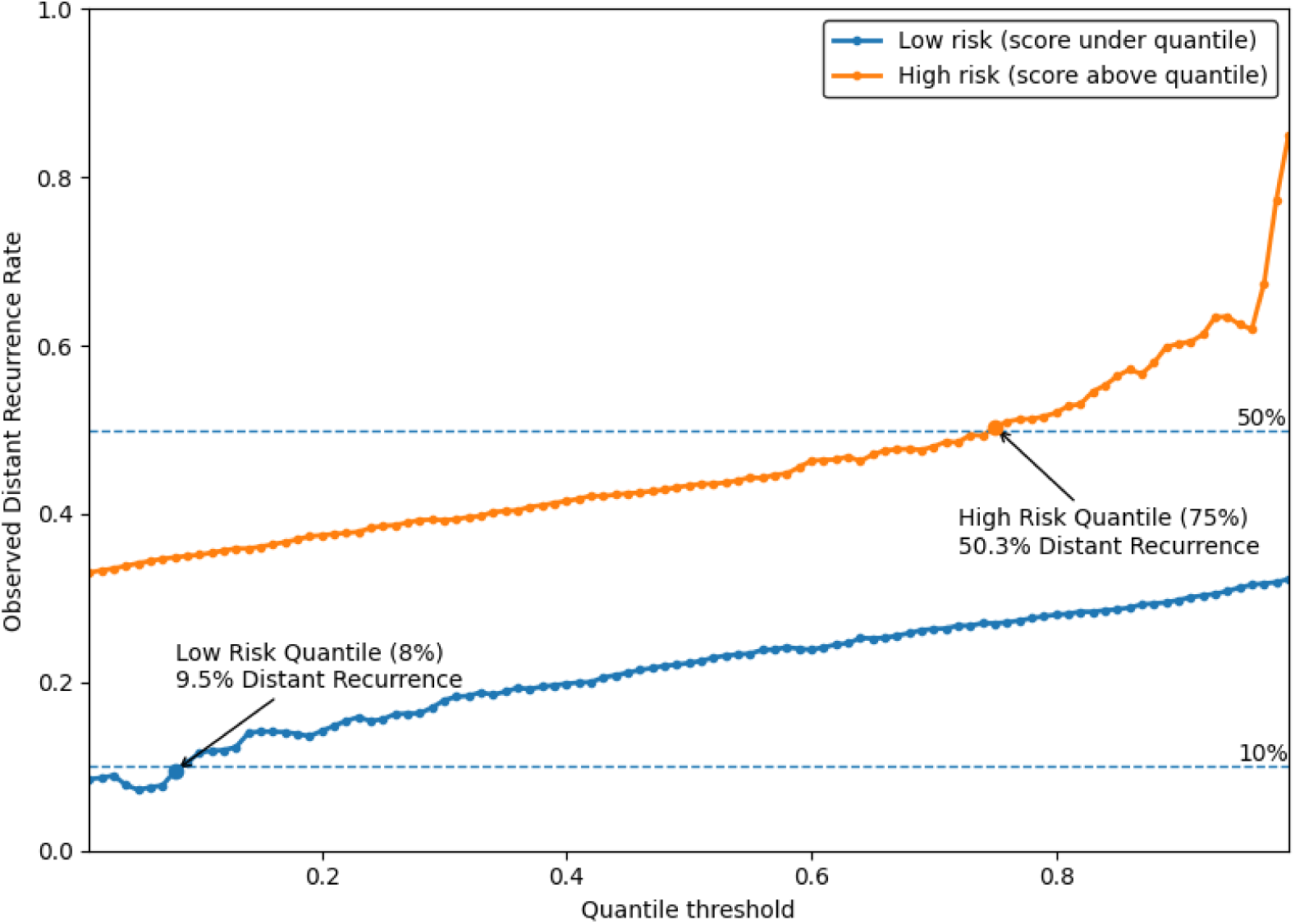
Determination of Clinically Relevant Risk Thresholds. Quantiles were assessed in 1% increments to identify thresholds defining low- and high-risk disease. A low-risk threshold was selected to identify patients in the CALGB 9344 cohort with a <10% risk of distant recurrence, while a high-risk threshold corresponded to >50% risk of distant recurrence. Risk quantiles demonstrated a generally monotonic relationship with recurrence risk, indicating that alternative thresholds could be chosen to stratify patients across a broad spectrum of risk relevant to adjuvant treatment decision-making. **Abbreviations:** CALGB – Cancer and Leukemia Group B.

**Supplemental Figure 4.**
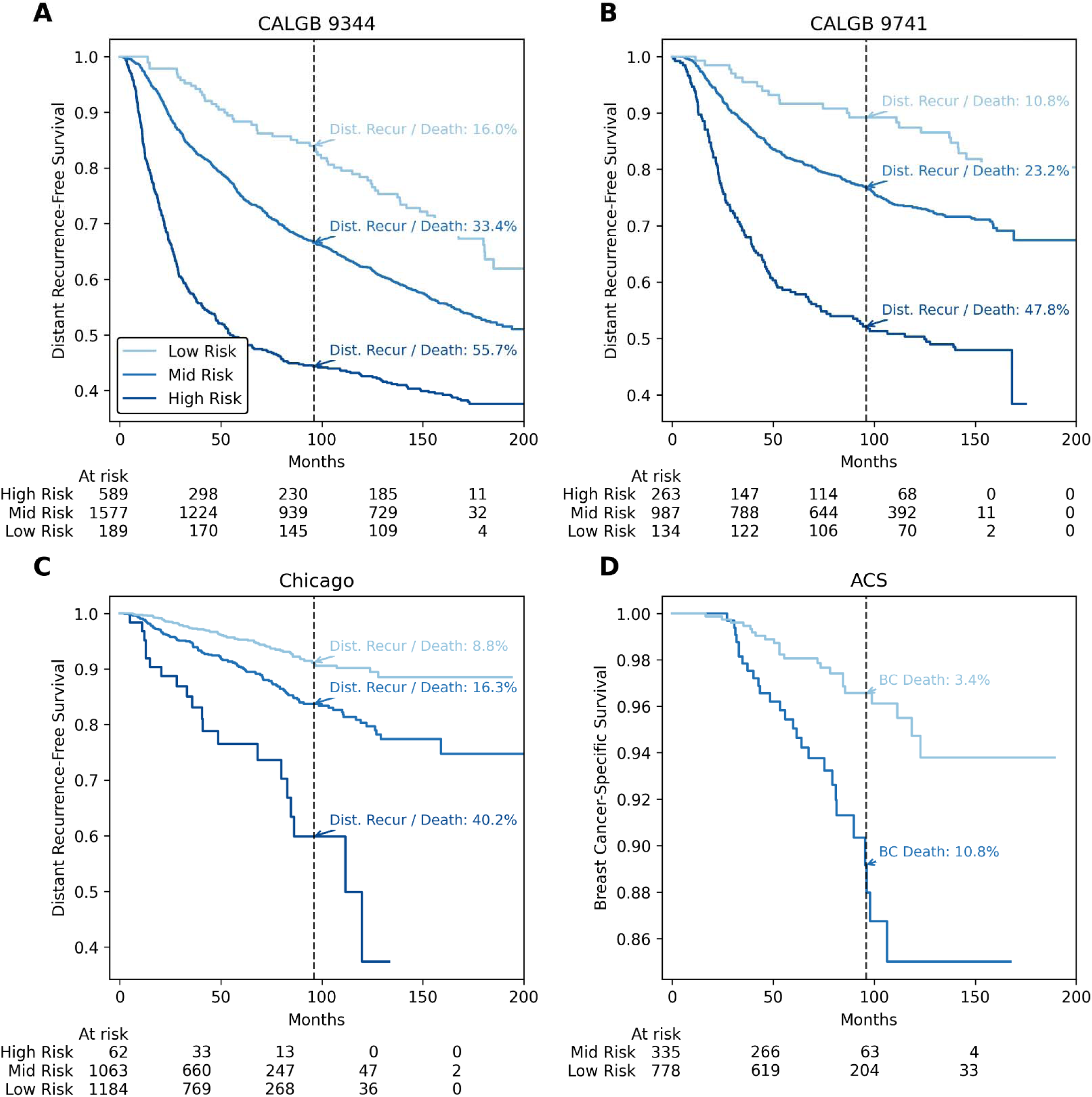
Multimodal Risk Stratification for Distant Recurrence-Free Survival Across Clinical Trial and Real-World Cohorts. Curves show low-, medium-, and high-risk groups defined by thresholds fit in CALGB 9344 to identify patients with a risk of distant recurrence below 10% (low risk) or above 50% (high risk). These predictions incorporate clinical covariates (ER status, PR status, tumor size, node status, age) and the histology-derived bronchioid signature. Shown are Kaplan-Meier estimates for distant-recurrence free survival in **A.** CALGB 9344; **B.** CALGB 9741; **C.** Chicago, and **D**. breast cancer-specific survival for ACS (as recurrence events were not captured in this cohort). The 8-year event rate is listed on each plot. **Abbreviations:** BC – Breast cancer; CALGB – Cancer and Leukemia Group B; ACS – American Cancer Society

**Supplemental Figure 5.**
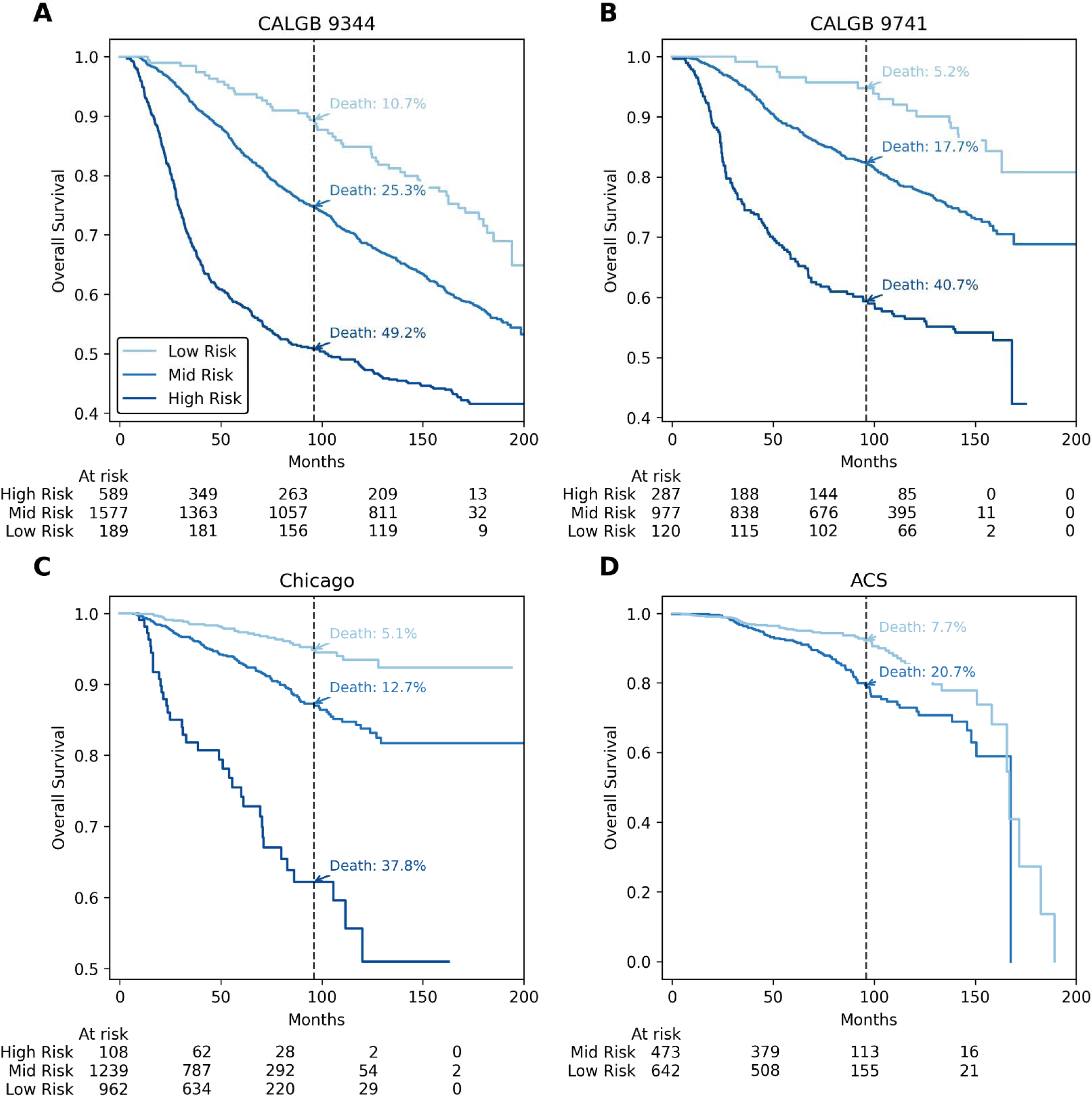
Multimodal Risk Stratification for Overall Survival Across Clinical Trial and Real-World Cohorts. Curves show low-, medium-, and high-risk groups defined by thresholds fit in CALGB 9344 to identify patients with a risk of distant recurrence below 10% (low risk) or above 50% (high risk). These predictions incorporate clinical covariates (ER status, PR status, tumor size, node status, age) and the histology-derived bronchioid signature. Shown are Kaplan-Meier estimates for overall survival in **A.** CALGB 9344; **B.** CALGB 9741; **C.** Chicago, and **D**. ACS cohorts. The 8-year event rate is listed on each plot. **Abbreviations:** CALGB – Cancer and Leukemia Group B; ACS – American Cancer Society

**Supplemental Figure 6.**
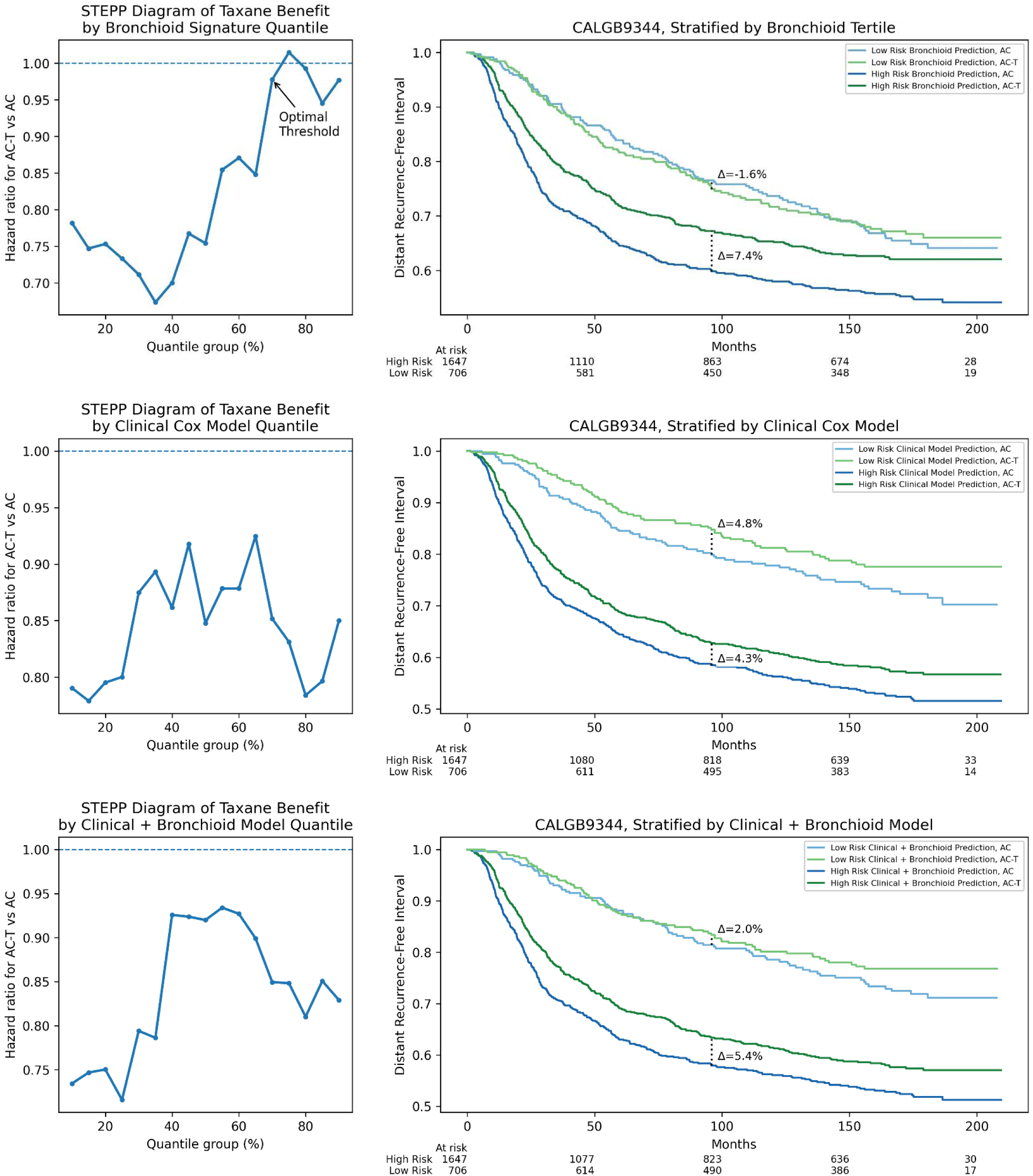
Histologic Risk More Accurately Predicts Chemotherapy Benefit than Clinical Risk. **Left:** Subpopulation treatment effect pattern plot illustrating the hazard ratio for taxane-containing chemotherapy (AC-T) versus AC in CALGB 9344 across overlapping quantile-defined subgroups of the bronchioid histology signature, a clinical Cox model fit on age, tumor size, nodal status, estrogen / progesterone status, and a clinicopathologic model also incorporating the bronchioid signature. **Right:** Kaplan-Meier estimates of distant recurrence-free interval in CALGB 9344 are shown, stratified by the 70^th^ percentile cutoff defined by **A.** the bronchioid signature, **B.** a clinical risk score from a Cox model fit on age, tumor size, nodal status, and estrogen / progesterone receptor status, and **C.** the multimodal risk derived from the signature and clinical variables. The 8-year landmark survival with annotated differences (Δ) in absolute survival between arms is listed **Abbreviations:** STEPP – Subpopulation Treatment Effect Pattern Plot; CALGB – Cancer and Leukemia Group B; AC – doxorubicin and cyclophosphamide; T – paclitaxel.

**Supplemental Figure 7.**
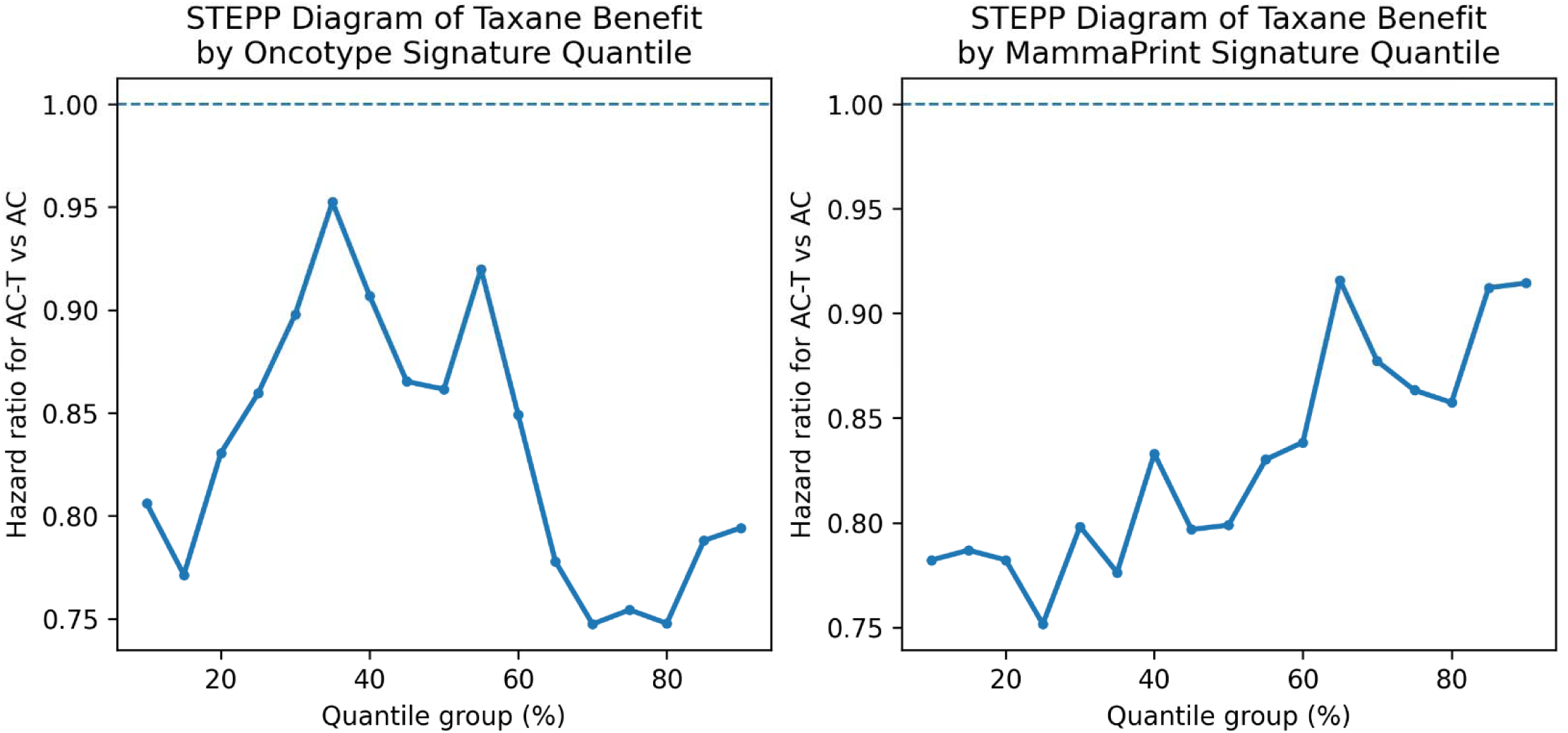
STEPP Diagrams for Taxane Benefit as a Function of Oncotype and MammaPrint Prediction. Subpopulation treatment effect pattern plot illustrating the hazard ratio for taxane-containing chemotherapy (AC-T) versus AC across overlapping quantile-defined subgroups of the Oncotype and MammaPrint histology signature scores in CALGB 9344. **Abbreviations:** STEPP – Subpopulation Treatment Effect Pattern Plot; CALGB – Cancer and Leukemia Group B; AC – doxorubicin and cyclophosphamide; T – paclitaxel.

**Supplemental Figure 8.**
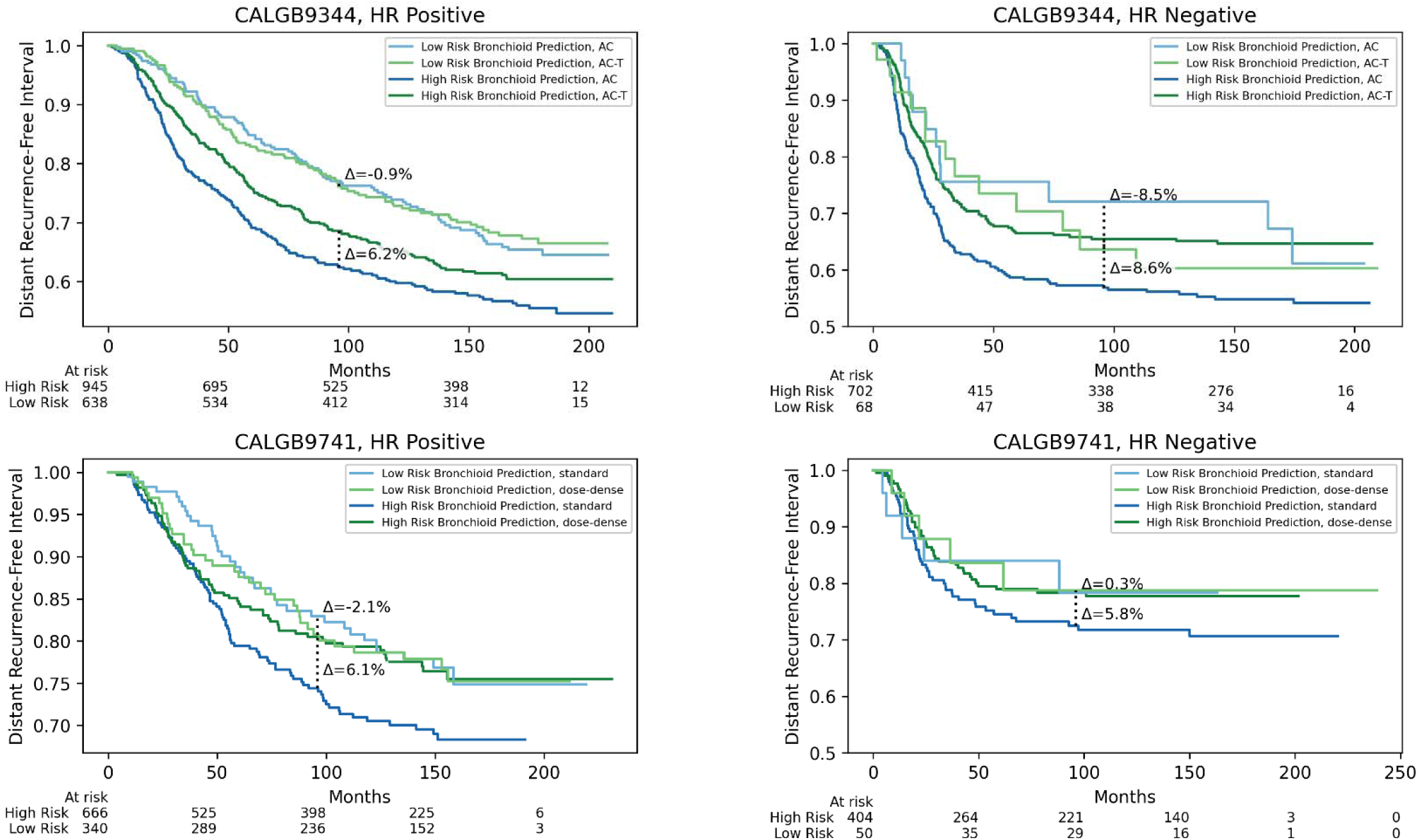
Predictive Value of the Bronchioid Histology Signature in Subgroups of Hormone Receptor Status. **Top:** Kaplan-Meier estimates of distant recurrence-free interval in CALGB 9344 in hormone receptor-positive and -negative subsets, stratified by predicted low- versus high-risk groups defined by the bronchioid signature (at the 70^th^ percentile) and randomized chemotherapy arm (AC vs AC-T). The 8-year landmark survival with annotated differences (Δ) in absolute survival between arms is listed. **Bottom:** Kaplan-Meier estimates of distant recurrence-free interval in CALGB 9741 in hormone receptor-positive and -negative subsets, comparing standard-schedule versus dose-dense chemotherapy. Curves are stratified by predicted risk group (low vs high) using the same signature threshold. The 8-year landmark survival with annotated differences (Δ) in absolute survival between arms is listed **Abbreviations:** CALGB – Cancer and Leukemia Group B; AC – doxorubicin and cyclophosphamide; T – paclitaxel; HR – hormone receptor.

**Supplemental Figure 9.**
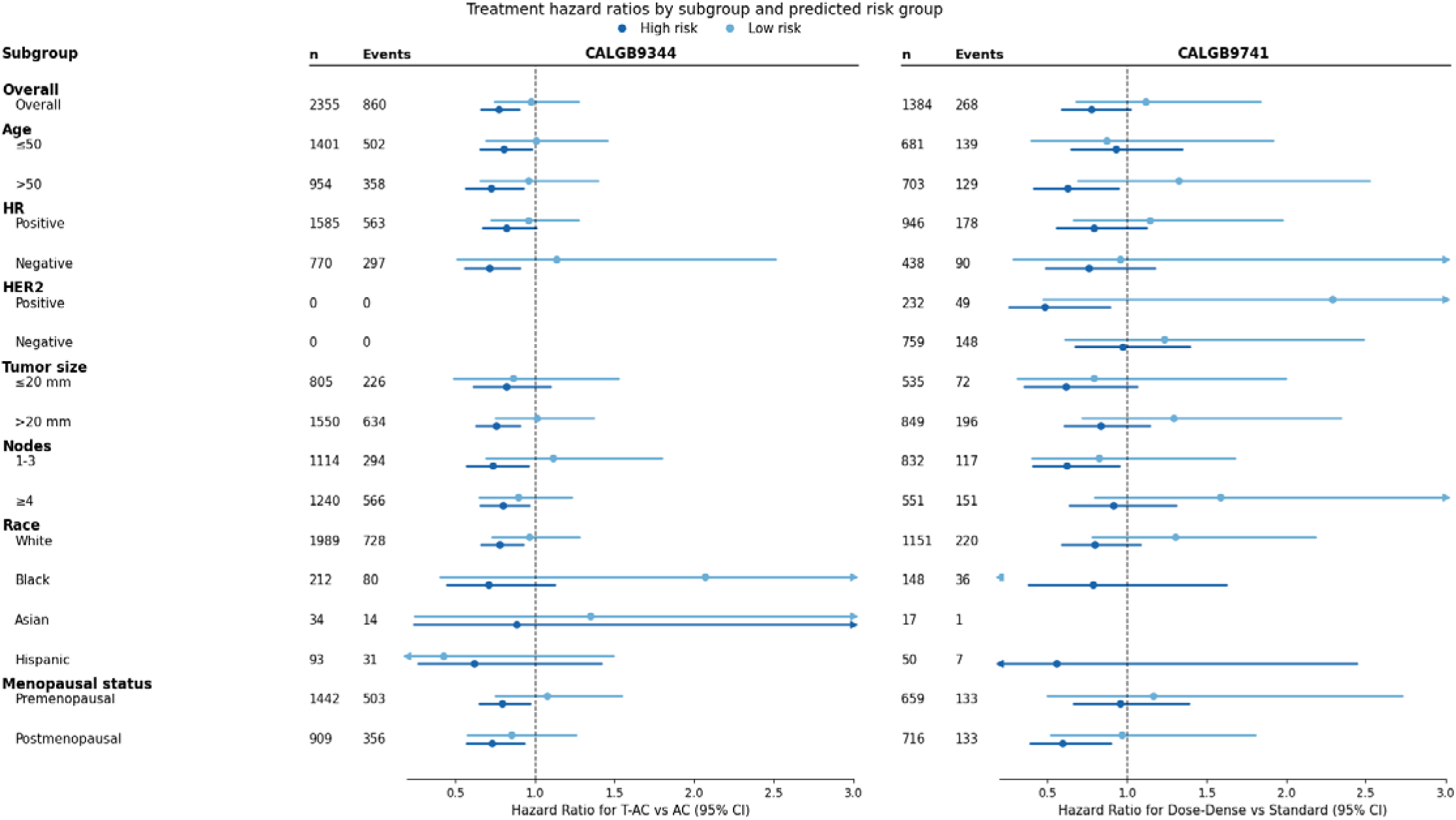
Treatment Effect by Predicted Risk Group Across Clinical Subgroups. Forest plots showing unadjusted hazard ratios and 95% confidence intervals for treatment effect (reduction in distant recurrence-free interval) stratified by bronchioid signature-defined low-risk and high-risk groups across clinicopathologic subgroups in CALGB 9344 and CALGB 9741 cohorts. **Abbreviations:** CALGB – Cancer and Leukemia Group B; AC – doxorubicin and cyclophosphamide; T – paclitaxel; HR – hormone receptor.

**Supplemental Table 1:** Grambsch-Therneau Proportional Hazards Test for Multivariable Cox model. Shown are per-covariate tests of the proportional hazards assumption based on scaled Schoenfeld residuals with the rank time transform for the CALGB 9344 dataset for a model trained to predict distant recurrence; the table reports the test χ² statistic and two-sided p-value. Restricting follow-up to five years reduced the violation of the proportional hazards assumption, especially for estrogen receptor status and bronchioid signature predictions.

|  | Across Full Dataset |  | Limited to < 5 Years |  |
| --- | --- | --- | --- | --- |
| Metric | Test-statistic | p | Test-statistic | p |
| Age | 0.153164 | 6.96E-01 | 0.000624 | 0.980073 |
| ER Status | 26.08826 | 3.26E-07 | 10.73908 | 0.001049 |
| # Nodes | 0.094201 | 7.59E-01 | 0.472776 | 0.491713 |
| PR Status | 7.455655 | 6.32E-03 | 8.469046 | 0.003612 |
| Tumor Size | 3.92421 | 4.76E-02 | 2.961119 | 0.085289 |
| Bronchioid Sig | 29.24935 | 6.36E-08 | 3.282779 | 0.07001 |

**Supplemental Table 2:** Accuracy for Recurrence Outcome Prediction from Bronchioid Path Signature, Clinical Factors, and Combination of Pathology and Clinical Factors. **Abbreviations:** CALGB – Cancer and Leukemia Group B; ACS – American Cancer Society; DRFI – Distant recurrence-free interval; DRFS – Distant recurrence-free survival; BCSS – Breast cancer-specific survival; OS – Overall survival.

| Dataset | Metric | Overall C-index |  |  | Early Recurrence (<5 Year) C-index |  |  |
| --- | --- | --- | --- | --- | --- | --- | --- |
|  |  | C-index, Path | C-index, Clin | C-index, Clin + Path | C-index, Path | C-index, Clin | C-index, Clin + Path |
| CALGB 9344 | DRFI | 0.587 | 0.642 | **0.654** | 0.625 | 0.672 | **0.690** |
|  | DRFS | 0.574 | 0.626 | **0.635** | 0.619 | 0.665 | **0.681** |
|  | OS | 0.587 | 0.640 | **0.647** | 0.647 | 0.715 | **0.727** |
| CALGB 9741 | DRFI | 0.561 | 0.613 | **0.626** | 0.598 | 0.661 | **0.676** |
|  | DRFS | 0.568 | 0.630 | **0.64** | 0.600 | 0.684 | **0.692** |
|  | OS | 0.579 | 0.660 | **0.662** | 0.620 | 0.731 | **0.733** |
| Chicago | DRFI | 0.682 | 0.743 | **0.769** | 0.698 | 0.739 | **0.781** |
|  | DRFS | 0.629 | 0.630 | **0.672** | 0.638 | 0.642 | **0.682** |
|  | OS | 0.632 | 0.701 | **0.709** | 0.642 | **0.718** | 0.717 |
| ACS | BCSS | 0.632 | 0.573 | **0.664** | 0.617 | 0.602 | **0.665** |
|  | OS | 0.557 | **0.689** | 0.641 | 0.526 | **0.699** | 0.621 |

**Supplemental Table 3:** Hazard Ratios for Outcome Prediction Across Low-, Medium-, and High-Risk Groups Defined by the Multimodal Model. * p < 0.05 **p < 0.001 **Abbreviations:** CALGB – Cancer and Leukemia Group B; ACS – American Cancer Society; DRFI – Distant recurrence-free interval; DRFS – Distant recurrence-free survival; BCSS – Breast cancer-specific survival; OS – Overall survival.

| Dataset | Metric | Hazard Ratio, (95%), p - value |  |  |
| --- | --- | --- | --- | --- |
|  |  | Low-Risk | Medium-Risk | High-Risk |
| CALGB 9344 | DRFI | ref | 2.52 (1.74 - 3.67)** | 5.11 (3.49 - 7.49)** |
|  | DRFS | ref | 1.59 (1.22 - 2.06)** | 2.98 (2.27 - 3.91)** |
|  | OS | ref | 1.72 (1.29 - 2.3)** | 3.31 (2.46 - 4.46)** |
| CALGB 9741 | DRFI | ref | 1.93 (1.21 - 3.09)* | 3.54 (2.16 - 5.82)** |
|  | DRFS | ref | 1.67 (1.15 - 2.43)* | 3.55 (2.39 - 5.26)** |
|  | OS | ref | 1.88 (1.19 - 2.96)* | 4.12 (2.58 - 6.60)** |
| Chicago | DRFI | ref | 4.76 (2.79 - 8.13)** | 18.31 (8.88 - 37.73)** |
|  | DRFS | ref | 2.11 (1.58 - 2.81)** | 6.22 (3.78 - 10.22)** |
|  | OS | ref | 2.67 (1.82 - 3.92)** | 10.2 (6.38 - 16.31)** |
| ACS | BCSS | ref | 2.96 (1.70 - 5.17)** | -- |
|  | OS | ref | 2.01 (1.44 - 2.81)** | -- |

**Supplemental Table 4:** Recurrence and Survival Outcomes in Low-, Medium-, and High-Risk Groups Identified by the Multimodal Clinicopathologic model. **Abbreviations:** CALGB – Cancer and Leukemia Group B; ACS – American Cancer Society; DRFI – Distant recurrence-free interval; DRFS – Distant recurrence-free survival; BCSS – Breast cancer-specific survival; OS – Overall survival

| Dataset | Metric | 4-Year Event Rate, % (95% CI) |  |  | 8-Year Event Rate, % (95% CI) |  |  |
| --- | --- | --- | --- | --- | --- | --- | --- |
|  |  | Low-Risk | Medium-Risk | High-Risk | Low-Risk | Medium-Risk | High-Risk |
| CALGB 9344 | DRFI | 4.4 (2.2 - 8.6) | 18.9 (17.0 - 20.9) | 43.3 (39.3 - 47.5) | 9.5 (6.0 - 14.8) | 29.1 (26.9 - 31.5) | 50.3 (46.2 - 54.5) |
|  | DRFS | 9.0 (5.7 - 14.1) | 20.4 (18.4 - 22.4) | 47.4 (43.5 - 51.6) | 16.0 (11.5 - 22.1) | 33.4 (31.1 - 35.9) | 55.7 (51.7 - 59.8) |
|  | OS | 3.7 (1.8 - 7.6) | 11.4 (10.0 - 13.1) | 38.8 (35.0 - 42.9) | 10.7 (7.0 - 16.1) | 25.3 (23.2 - 27.5) | 49.2 (45.2 - 53.4) |
| CALGB 9741 | DRFI | 4.2 (1.9 - 9.1) | 13.7 (11.7 - 15.9) | 27.8 (22.7 - 33.9) | 10.3 (6.2 - 16.7) | 20.7 (18.2 - 23.3) | 34.9 (29.1 - 41.4) |
|  | DRFS | 7.2 (3.9 - 12.9) | 16.1 (14.0 - 18.5) | 38.0 (32.4 - 44.1) | 14.8 (9.8 - 22.0) | 26.0 (23.4 - 28.8) | 49.4 (43.5 - 55.6) |
|  | OS | 1.6 (0.4 - 6.2) | 8.8 (7.2 - 10.7) | 28.7 (23.9 - 34.2) | 5.8 (2.8 - 11.8) | 18.6 (16.3 - 21.2) | 40.7 (35.3 - 46.7) |
| Chicago | DRFI | 1.0 (0.6 - 1.9) | 4.3 (3.2 - 5.9) | 20.2 (11.6 - 33.8) | 1.9 (1.1 - 3.2) | 8.8 (6.8 - 11.4) | 25.8 (15.5 - 40.9) |
|  | DRFS | 3.3 (2.4 - 4.6) | 7.6 (6.0 - 9.5) | 21.2 (12.5 - 34.5) | 8.8 (6.9 - 11.3) | 16.3 (13.6 - 19.4) | 40.2 (26.5 - 57.6) |
|  | OS | 1.7 (1.0 - 2.8) | 5.3 (4.1 - 6.9) | 19.3 (12.9 - 28.4) | 5.1 (3.5 - 7.4) | 12.7 (10.5 - 15.5) | 37.8 (28.2 - 49.5) |
| ACS | BCSS | 1.1 (0.6 - 2.2) | 3.5 (1.9 - 6.2) | -- | 3.4 (2.2 - 5.4) | 10.8 (7.0 - 16.5) | -- |
|  | OS | 3.6 (2.4 - 5.4) | 6.4 (4.5 - 9.1) | -- | 7.7 (5.5 - 10.8) | 20.7 (16.3 - 26.0) | -- |

**Supplemental Table 5:** Adjusted Treatment Effects in Low- versus High-Risk Groups Defined by the Bronchioid Signature. Low-risk is defined as cases with bronchioid signature predictions at or above the 70^th^ percentile cutoff from CALGB 9344. Hazard ratios indicate the survival benefit associated with treatment intensification (AC-T vs AC for CALGB 9344; dose-dense vs standard-schedule chemotherapy for CALGB 9741; chemotherapy vs none for Chicago), and are adjusted for age, tumor size, nodal status, estrogen / progesterone status, and signature prediction. The interaction hazard ratio is calculated for the same full parameterized model, with p-value given for likelihood ratio test for model with and without interaction term. **Abbreviations:** CALGB – Cancer and Leukemia Group B; HR – hazard ratio; DRFI – Distant recurrence-free interval; DRFS – Distant recurrence-free survival; OS – Overall survival; AC – doxorubicin and cyclophosphamide; T – paclitaxel.

| Dataset | Metric | HR, high-risk | p | HR, low-risk | p | Interaction HR | p |
| --- | --- | --- | --- | --- | --- | --- | --- |
| **CALGB 9344**<br>*AC-T vs AC* | DRFI | 0.78 (0.67 - 0.91) | 0.00206 | 1.00 (0.76 - 1.30) | 0.975 | 1.26 (0.93 - 1.72) | 0.136 |
|  | DRFS | 0.76 (0.66 - 0.88) | 0.000141 | 1.02 (0.81 - 1.27) | 0.883 | 1.34 (1.03 - 1.75) | 0.0284 |
|  | OS | 0.77 (0.66 - 0.89) | 0.000492 | 1.02 (0.80 - 1.31) | 0.86 | 1.34 (1.01 - 1.78) | 0.0426 |
| **CALGB 9741**<br>*Dose-dense vs Standard* | DRFI | 0.74 (0.58 - 0.95) | 0.0165 | 1.12 (0.73 - 1.71) | 0.603 | 1.48 (0.91 - 2.39) | 0.123 |
|  | DRFS | 0.69 (0.56 - 0.85) | 0.000463 | 1.09 (0.76 - 1.57) | 0.633 | 1.55 (1.03 - 2.34) | 0.0391 |
|  | OS | 0.71 (0.57 - 0.89) | 0.00279 | 1.09 (0.73 - 1.64) | 0.673 | 1.51 (0.95 - 2.40) | 0.0785 |
| **Chicago**<br>*Chemo vs No Chemo* | DRFI | 1.15 (0.67 - 2.00) | 0.609 | 0.58 (0.0 - 5633484.46) | 0.946 | 14.66 (0.60 - 359.58) | 1 |
|  | DRFS | 0.84 (0.59 - 1.21) | 0.361 | 1.42 (0.90 - 2.26) | 0.135 | 1.91 (1.17 - 3.14) | 0.00866 |
|  | OS | 0.85 (0.57 - 1.27) | 0.42 | 1.11 (0.65 - 1.90) | 0.694 | 1.51 (0.87 - 2.62) | 0.134 |

